# Hydroxychloroquine plus standard care compared with the standard care alone in COVID-19: a meta-analysis of randomized controlled trials

**DOI:** 10.1101/2020.06.05.20122705

**Authors:** Bahman Amani, Ahmad Khanijahani, Behnam Amani

**Affiliations:** School of Public Health, Tehran University of Medical Sciences, Poursina St, Ghods St. Tehran, Iran; School of Health Sciences, Duquesne University, 600 Forbes Ave, Pittsburgh, PA 15282

**Author notes:** **Corresponding Author:** Behnam Amani.

**Keywords:** Coronavirus Disease, COVID-19, SARS-CoV-2, Hydroxychloroquine, Efficacy, Safety

## Abstract

**Background & Objective:** The efficacy and safety of Hydroxychloroquine (HCQ) in treating coronavirus disease COVID-19 pandemic is disputed. This study aimed to examine the efficacy and safety of HCQ plus the standard of care in COVID-19 patients.

**Methods:** PubMed, The Cochrane Library, Embase, and web of sciences were searched up to June 1, 2020. The references list of the key studies was reviewed for additional relevant resources. Clinical studies registry databases were searched for identifying potential clinical trials. The quality of the included studies was evaluated using the Cochrane Collaboration’s tool. Meta-analysis was performed using RevMan software (version 5.3).

**Results:** Three randomized controlled trials with total number of 242 patients were identified eligible for meta-analysis. No significant differences were observed between HCQ and standard care in terms of viral clearance (Risk ratio [RR] = 1.03; 95% confidence interval [CI] = 0.91, 1.16; P = 0.68), disease progression (RR = 0.92; 95% CI = 0.10, 0.81; P = 0.94), Chest CT (RR = 1.40; 95% CI = 1.03, 1.91; P = 0.03). There is a significant difference between HCQ and standard care for adverse events (RR = 2.88; 95% CI = 1.50, 5.54; P = 0.002).

**Conclusion:** Although the current meta-analysis failed to confirm the efficacy and safety of HCQ in the treatment of COVID-19 patients, further rigorous randomized clinical trials are necessary to evaluate conclusively the efficacy and safety of HCQ against COVID-19.

## BACKGROUND

The epidemic of coronavirus disease (COVID-19) in late December 2019, originated from the city of Wuhan in China and became pandemic [1,2].

COVID-19 is the third most common coronavirus infection epidemy in the last twenty years after diseases of acute respiratory syndrome (SARS) and the Middle East respiratory syndrome (MERS) [3]. The mortality of critically ill patients with COVID-19 is considerable [4]. The initial estimations for case fatality rate is about 2.3% in China [5] and 7.2% in Italy [6]. The 2019-nCoV infection causes clusters of severe respiratory illness similar to severe acute respiratory syndrome (SARS) coronavirus and, in severe cases, was associated with hospitalization, ICU admission, and frequent mortalities [7,8]. Fever, cough, shortness of breath, muscle ache, confusion, headache, sore throat, rhinorrhea, chest pain, diarrhea, nausea, and vomiting are among the clinical manifestation of the disease [9]. Early efforts have focused on describing the clinical course, containing severe cases, and treating the sick [2]. Currently, there is no drug or vaccine approved for the treatment of human coronavirus [4].

There are several options for the control and preventing the development of COVID-19 infections including vaccines, monoclonal antibodies, oligonucleotide-based therapies, peptides, interferon therapies, and small-molecule drugs [4]. Lopinavir/Ritonavir (400/100 mg every 12 hours), Chloroquine (500 mg every 12 hours), and Hydroxychloroquine (HCQ) (200 mg every 12 hours) and Alpha-interferon are also proposed as courses of treatment [3].

HCQ is used for a variety of diseases including IgAN [10,11], Arthritis [12,13], Pulmonary Sarcoidosis [14], Cutaneous lupus erythematosus [15–17], Sjogren’s syndrome [18] and Type 2 diabetes mellitus [19]. Currently several observational [20,21]and clinical studies [22,23] have evaluated the efficacy of HCQ on COVID-19.

Therefore, there is an urgent need to examine and conclude on this issue based on available evidence. The purpose of this study was to examine the efficacy and safety of HCQ in COVID-19 patients.

## METHODS

The study protocol was registered in the international prospective reregister of systematic review (PROSPERO) with the registration number of CRD42020179425. We used the preferred reporting items for systematic reviews and meta-analyses (PRISMA) checklist when writing this report [24].

### Search strategy

A systematic review of the relevant literature was conducted in PubMed, Cochrane Library, Embase and Google scholar up to June 1, 2020. The references list of the final studies and review articles were reviewed for more citations. The European Union Clinical Trials Register, Clinical trial Gov. and Chinese Clinical Trial Registry (ChiCTR) were also searched. Search terms included 2019-nCoV, SARS-CoV-2, COVID-19, and Hydroxychloroquine which were usually limited to title and abstract of the articles in our search strategy. There was no restriction on the language.

### Study selection

Two researchers were independently screened the identified studies based on the inclusion criteria. Disagreements were resolved by discussion among the authors. The following inclusion criteria were used for selecting the articles: (1) Patients with confirmed COVID-19; (2) HCQ as intervention; (3) Standard treatment as control; (4) Randomized controlled trial as study design; (5) and clinical recovery, mortality rate or any adverse events as an outcome. Other studies and reports such as letter to the editor, case reports, editorial comments, observational studies and animal models were excluded

### Data Extraction and Quality Assessment

Cochrane Collaboration’s tool was used to assess the risk of bias in the selected studies. Information on the study characteristics (place, design, and duration), patient’s characteristics (age, sex, and number of patients), intervention and control (treatment protocol), efficacy outcomes (clinical improvement and mortality rate), and adverse events were extracted. Both processes were independently performed by two authors, and disagreements were resolved by discussion among the authors

### Quantitative data synthesis

A meta-analysis was conducted to compare the efficacy and safety of HCQ versus standard treatment using RevMan (version 5.3) software. For dichotomous variables, risk ratio (RR) and a 95% confidence interval (CI) were used. Statistical heterogeneity was evaluated using the I2 and Chi-square tests with a significance level p < 0.05. The random-effects and fixed-effect methods were used for studies with heterogeneity high and low to moderate and, respectively.

## RESULTS

### The process of selecting studies

Figure. 1 shows the process of literature search, removal of duplication, and screening based. Of a total of 97 records, 91 were excluded based on the title and abstract. Three studies involving 242 patients [22,23,25] were included in the meta-analysis.

**Figure 1.**
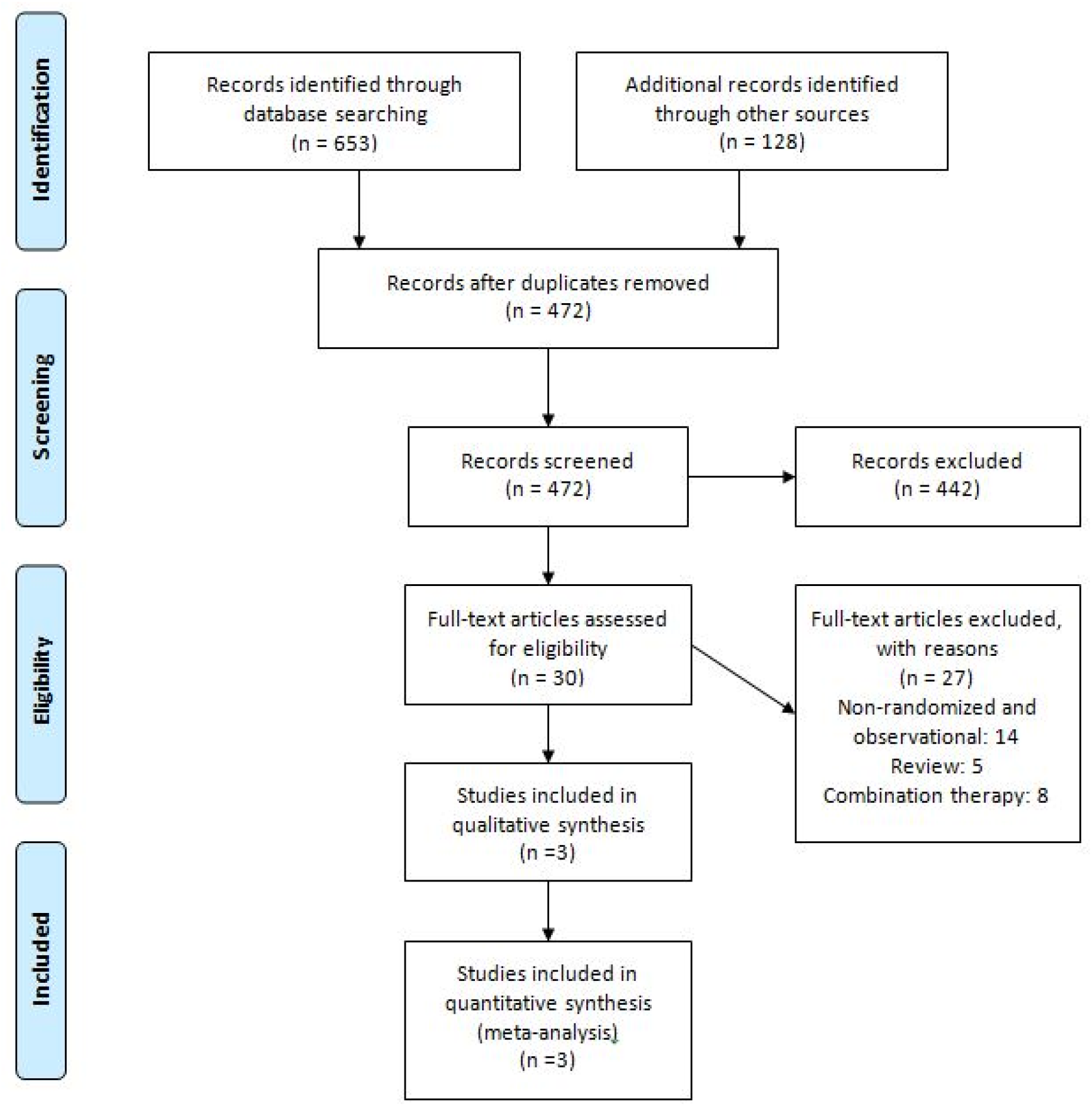
Flowchart of study selection process

### Study characteristics

The main characteristics of selected are presented in Table. 1.

**Table 1.** The characteristics of RCTs included in the meta-analysis

| First author, year, country | Patients (N, Females, Mean Age) | Intervention (N, HCQ protocol) | Control (N, protocol) | Duration (days) | Inclusion criteria | Outcomes |
| --- | --- | --- | --- | --- | --- | --- |
| Chen, 2020, China | 62, 33, 44.7 | 33; (400 mg/d (200 mg/bid) between days 1 and 5) + Control treatment | 29; Oxygen therapy, antiviral agents, antibacterial agents, and immunoglobulin, with or without corticosteroids (standard treatment) | 5 | (1) Age $\geq 18$ years; (2) Laboratory (RT-PCR) positive of SARS-CoV-2; (3) Chest CT with pneumonia; (4) SaO<sub>2</sub>/SPO<sub>2</sub> ratio > 93% or PaO<sub>2</sub>/FIO<sub>2</sub> ratio > 300 mmHg under the condition in the hospital room (mild illness) | (1) TTCR (defined as the return of body temperature and cough relief); (2) Radiological changes; (3) Pulmonary recovery |
| Chen J, 2020, China | 30, 9, 48.6 | 15; (400 mg per day for 5 days) + Control treatment | 15; bed rest, oxygen inhalation, symptomatic support therapy, using the anti-treatment recommended in the "diagnosis and treatment plan" Viral drugs such as alpha interferon nebulization, oral lopinavir/ritonavir | 7 | Age $\geq 18$ years old; confirmed COVID-19 | Negative conversion rate of COVID-19 nucleic acid in respiratory pharyngeal swab on days 7 |
| Tang, 2020, China | 150, 68, 46.1 | 75; HCQ 200 mg/d x 3 d followed by 800/d + Standard care | 75; Standard care | 14-21 | (1) age > 18 years; (2) confirmed COVID-19; | RT-PCR negativity on d 28, clinical progression, adverse events |

Abbreviations: TTCR: Changes in time to clinical recovery; N: Sample size; HCQ: Hydroxychloroquine

Chen and et al [22] conducted a randomized clinical trial to examine the efficacy of HCQ in patients with confirmed COVID-19 in China. 62 patients were randomly assigned into two groups. According to the findings, HCQ resulted in significant improvement in the HCQ treatment group versus the control group (80.6% vs. 54.8%, P < 0.05). There was a significant difference in fever and cough between two groups (P < 0.05).

Tang et al [23] examined the efficacy and safety of HCQ patients with confirmed COVID-19. They found that HCQ did not result in clinical improvement of COVID-19 patients. At the endpoint, 85.4% of HCQ –treated patients were cured compared to 81.3% in the control group.

Chen J et al [25] evaluated the efficacy and safety of HCQ patients with COVID-19 in China. Thirty patients were randomly assigned to HCQ and control groups. At the end of the study, 86.7% of patients in the HCQ group and 93.3% of patients in the control group were cured (P 0.05). Quality assessment of the studies and risk of bias, using the Cochrane Collaboration’s tool, is presented in Figure. 2

**Figure 2.**
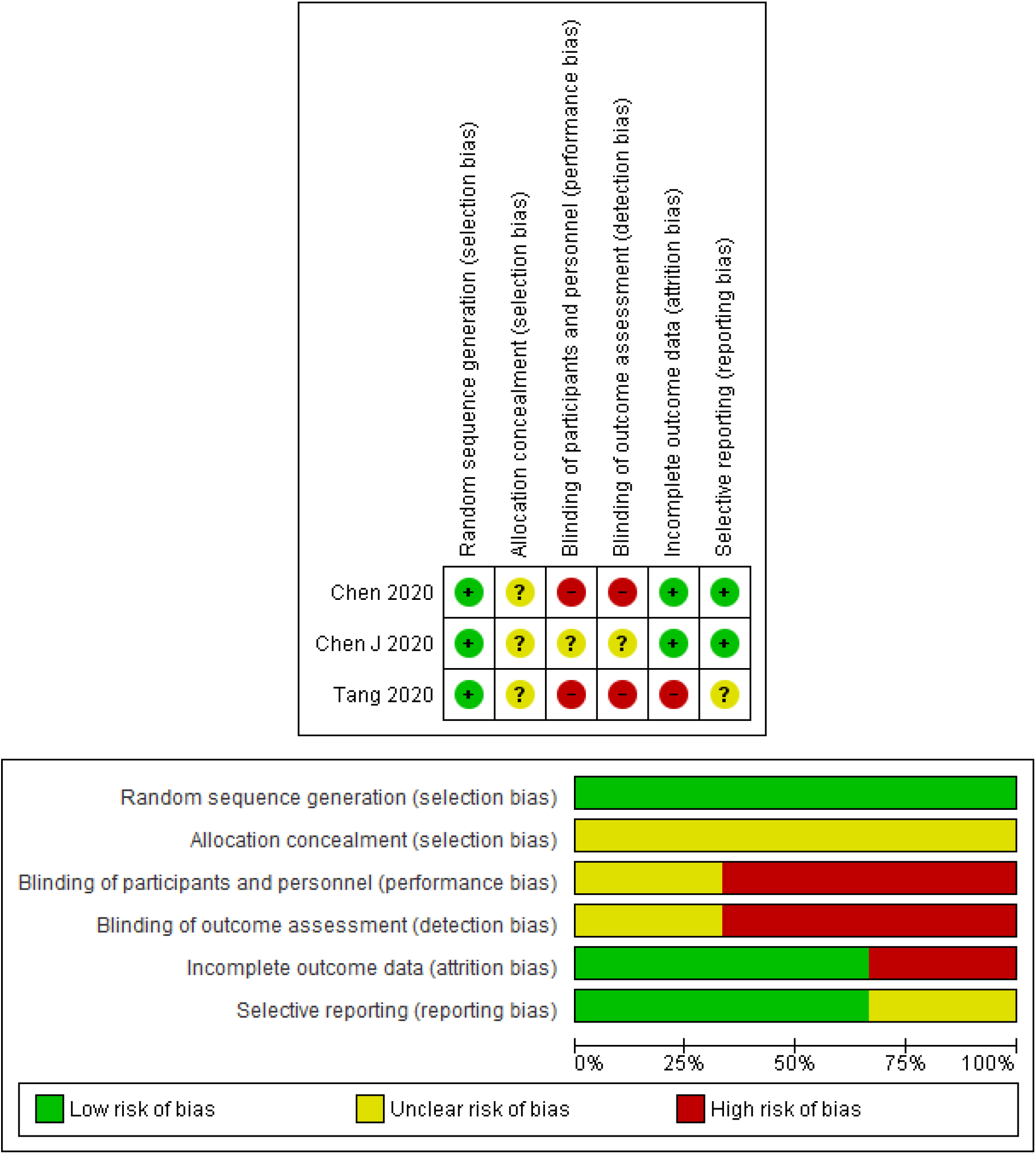
Risk of bias in the selected studies

### Efficacy

Viral clearance: pooled data of two studies [23,25] demonstrated that there is no significant difference between HCQ and control groups (RR = 1.03; 95% confidence interval [CI] = 0.91, 1.16; P = 0.68).(Figure 3. Section A).

Disease progression: pooled data of two studies [22,23,25] demonstrated that there is no significant difference between HCQ and control groups (RR = 0.92; 95% CI = 0.10, 0.81; P = 0.94).(Figure 3. Section B).

**Figure 3.**
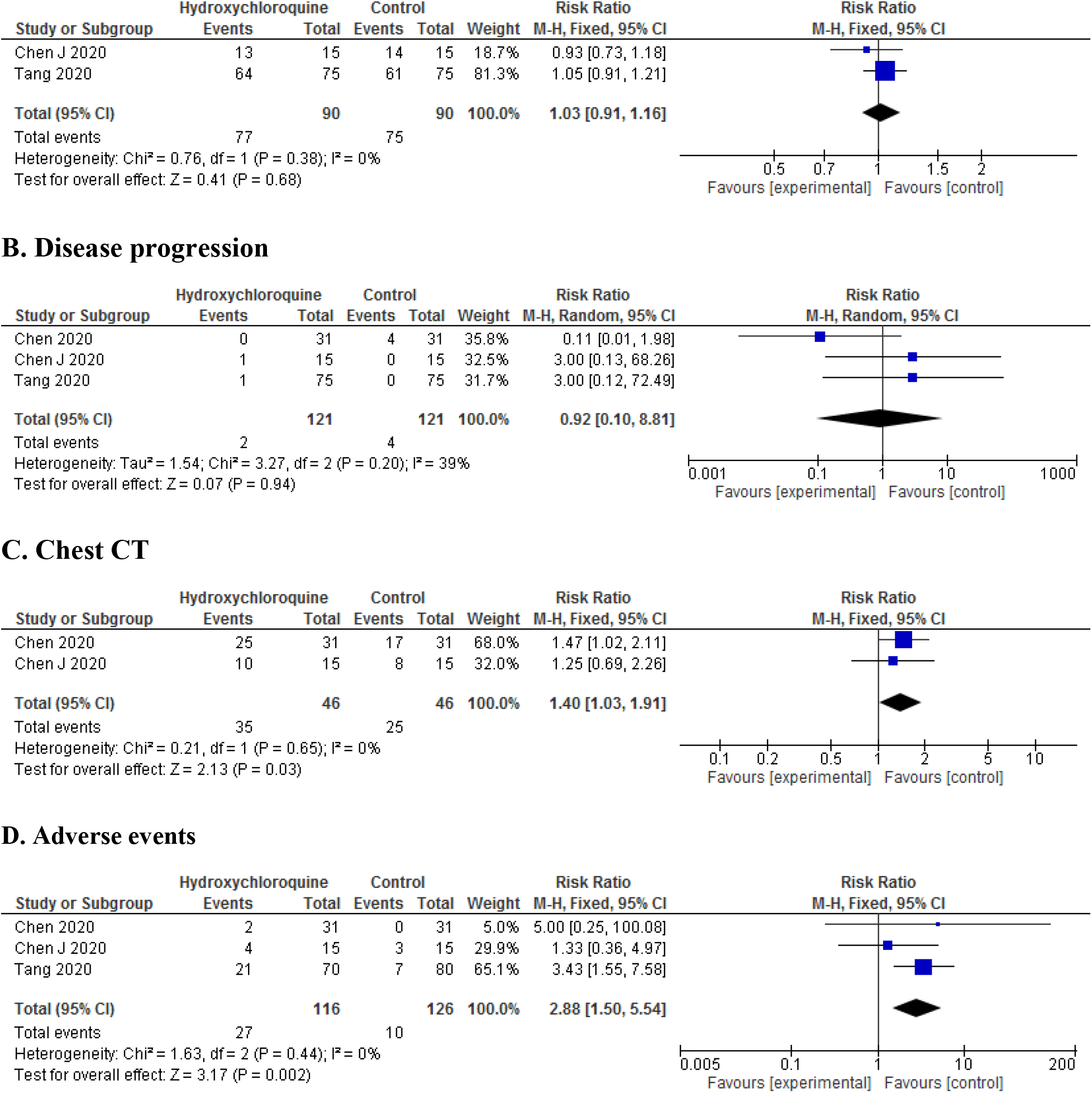
Forest plot of HCQ versus control, (A) Viral clearance, (B) Disease progression, (C) Chest CT, (D) Adverse events comparing HCQ and control

Chest CT: pooled data of two studies [22,25] demonstrated that there is no significant difference between HCQ and control groups (RR = 1.40; 95% CI = 1.03, 1.91; P = 0.03). (Figure 3. Section C).

### Adverse events

Adverse events were observed in three studies [22,23,25]. In the Chen et al [22] study, in two patients (6.4%) in HCQ group rash and headache were observed, while in the control group there were no adverse events. In a pilot study, four (26.7%) patients in the HCQ group and three (20%) patients in the control group were reported diarrhea and abnormal liver function (P>0.05). In Tang et al [23] study, adverse events were observed in 21 (30%) patients HCQ and 7 (8.8%) HCQ non-recipients (Figure 3. Section D). Pooled data demonstrated that there is a significant difference between HCQ and standard care for adverse events (RR = 2.88; 95% CI = 1.50, 5.54; P = 0.002). (Figure 3. Section D)

[Figure 3. goes here]

## DISCUSSION

The purpose of this study was to examine the efficacy and safety of HCQ compared to standard intervention for COVID-19. Although there is no currently proven treatment for COVID-19, several treatment strategies have been suggested for this condition.

A few RCTs examined the efficacy and safety of HCQ in treating COVID-19. The findings of our meta-analysis showed that there is no significant difference between HCQ and standard care in patients with confirmed COVID-19 (P>0.05). Previously published meta-analyses [26–28] on observational studies and randomized controlled trials found no clinical benefits for HCQ in comparison with standard care for COVID-19 patients. These studies confirm our findings.

It should be noted that the most of included studies in these meta-analyses were observational. There are some concerns regarding the limitations of these studies which should be considered. All kinds of biases such as confounding, reverse causation, statistical considerations and other issues are the limitations of these studies in the estimation of drug efficacy and safety [29]. The Agency for Healthcare Research and Quality (AHRQ) has provided recommendations on including observational studies into the comparative effectiveness review process for comparing medical interventions [30].

Gautret et al [31] suggested that the causes of insufficient response to treatment with HCQ in the non-respondents with COVID-19 should be examined by factors such as SARS-CoV-2 strains, genome, and other factors associated with the metabolism of HCQ in patients. A possible mechanism for HCQ inefficiency was explained by Sandeep and McGregor [32] using virtualized quantum mechanical modeling. However, Yao et al [33] found that HCQ was more potent than Chloroquine inhabiting SARS-CoV-2 in vitro.

Generally, the evidence for the efficacy and safety of HCQ is controversial. A meta-analysis by Shamshirian et al [28] confirms these findings and showed that HCQ treatment is associated with higher mortality (1.5 times). However, another Meta-analysis study showed that there was no significant difference between HCQ and control treatment for mortality [26].

Some studies supported the synergistic effect of HCQ with Azithromycin on COVID-19. A study [34] demonstrated the moderate in vitro effect of HCQ alone and its synergistic effect as combination therapy with Azithromycin on COVID-19. In an open-label non-randomized clinical trial, Gautret et al [31] found that 100% of patients received HCQ and Azithromycin as combination therapy were cured. These authors found similar results in another study. A meta-analysis showed that HCQ alone or as the combination with AZM in comparison with the control group was not effective in COVID-19 and was associated with higher mortality rates [35].

A newly published meta-analysis showed that not only the combination of HCQ and azithromycin did not result in any significant clinical improvement in COVID-19 patients, but also it was associated with 2.5 times higher mortality in comparison to the control group. The mortality was 1.5 times more compared to solo HCQ treatment. Contradictorily, a study on 1438 patients hospitalized with Covid-19 in all United States Veterans Health Administration medical centers found that the rates of death in patients under treatment with HCQ alone were higher than and HCQ + Azithromycin (27.8%, 22.1%) [21].

A meta-analysis of the adverse effects of long-term Azithromycin use in patients with chronic lung diseases showed that Azithromycin is associated with increasing the risk of bacterial resistance (2.7-fold) [36]. Our meta-analysis showed that there is a significant difference between HCQ and the control group for adverse events. Reported adverse events in RCTs were mild. Adverse effects of antimalarial are usually rare and mild [37]. Gastric symptoms are a prevalent adverse effect of HCQ [38,39], Other adverse events are including Cutaneous [40], headache [41–43], Cardiomyopathy [44–46] and Retinopathy [47–51]. Regarding the short-term follow-up of the studies, it is recommended that patients received HCQ should be monitored for possible adverse events for a longer period.

### Limitations

The existence of a limited number of studies, high heterogeneity between studies, small sample size, short follow-up period, and lack of rigor methodology of the studies were among the limitations of our study. Nevertheless, the findings of this study can be beneficial for guiding clinicians in decisions regarding COVID-19 treatment.

## CONCLUSION

According to the findings of this study, HCQ appears to be not effective and safe in the treatment of patients with COVID-19. Given that there are limited RCTs on the efficacy and safety of HCQ in COVID-19, there are needed more evidence. Further well-designed and high quality randomized controlled trials with large sample size are necessary to establish the efficacy and safety of HCQ for this disease.

## Data Availability

The data that support the findings of this study are openly available at:
Chen, Z. et al. Efficacy of hydroxychloroquine in patients with COVID-19: results of a randomized clinical trial. MedRxiv (2020).
Tang, W. et al. Hydroxychloroquine in patients with COVID-19: an open-label, randomized, controlled trial. MedRxiv (2020).
Chen, J. et al. A pilot study of hydroxychloroquine in treatment of patients with common coronavirus disease-19 (COVID-19). Journal of Zhejiang University (Medical Science) 49, 0-0 (2020).

## Acknowledgments

We are thankful to the authors of the studies included in this systematic review and meta-analysis.

## Author Contribution

BaA and BeA systematically reviewed the search results and selected the relevant studies. BaA performed the formal analysis. All authors performed the literature search, wrote the manuscript, and reviewed the manuscript.

